# Use cases for COVID-19 screening and surveillance with rapid antigen-detecting tests: a systematic review

**DOI:** 10.1101/2021.11.03.21265807

**Authors:** Apoorva Anand, Jacob Bigio, Emily MacLean, Talya Underwood, Nitika Pant Pai, Sergio Carmona, Samuel G. Schumacher, Amy Toporowski

## Abstract

**Introduction:** Testing is critical to controlling the COVID-19 pandemic. Antigen-detecting rapid diagnostic tests (Ag-RDTs) that can be used at the point of care have the potential to increase access to COVID-19 testing, particularly in settings with limited laboratory capacity. This systematic review synthesized literature on specific use cases and performance of Ag-RDTs for detecting SARS-CoV-2, for the first comprehensive assessment of Ag-RDT use in real-world settings.

**Methods:** We searched three databases (PubMed, EMBASE and medRxiv) up to 12 April 2021 for publications on Ag-RDT use for large-scale screening, irrespective of symptoms, and surveillance of COVID-19, excluding studies of only presumptive COVID-19 patients. We tabulated data on the study setting, populations, type of test, diagnostic performance and operational findings. We assessed risk of bias using QUADAS-2 and an adapted tool for prevalence studies.

**Results:** From 4313 citations, 39 studies conducted in asymptomatic and symptomatic adults were included. Study sample sizes varied from 40 to >5 million. Of 39 studies, 37 (94.9%) investigated lateral flow Ag-RDTs and two (5.1%) investigated multiplex sandwich chemiluminescent enzyme immunoassay Ag-RDTs. Six categories of testing (screening/surveillance) initiatives were identified: mass screening (n=13), targeted screening (n=11), healthcare entry testing (n=6), at-home testing (n=4), surveillance (n=4) and prevalence survey (n=1). Across studies, Ag-RDT sensitivity varied from 40% to 100%. Ag-RDTs were noted as convenient, easy-to-use and low cost, with a rapid turnaround time and high user acceptability. Risk of bias was generally low or unclear across the studies.

**Conclusion:** This systematic review demonstrates the use of Ag-RDTs across a wide range of real-world settings for screening and surveillance of COVID-19 in both symptomatic and asymptomatic individuals. Ag-RDTs were overall found to be easy-to-use, low cost and rapid tools, when consideration is given to their implementation and interpretation. The review was funded by FIND, the global alliance for diagnostics.

**SUMMARY:** *What is already known?:*

- Antigen-detecting rapid diagnostic tests (Ag-RDTs) have the potential to substantially improve access to timely testing for COVID-19 and are being deployed in a variety of settings around the world
- While studies have investigated the diagnostic accuracy of Ag-RDTs, less is known about how and in what settings Ag-RDTs are being used around the world and their performance in these different settings

*What are the new findings?:*

- Ag-RDTs are being used in a diverse range of real-world settings for mass screening and surveillance of COVID-19 among symptomatic and asymptomatic individuals
- The sensitivity of Ag-RDTs is variable - ranging from 40% to 100% - and in some cases low compared with RT-PCR, meaning that the value of testing with Ag-RDTs needs to be carefully evaluated for each use case taking into account factors such as the prevalence of COVID-19 in the population, the consequences of false positive or false negative results, and whether confirmatory testing of positive or negative Ag-RDT results with RT-PCR is required
- Nevertheless, Ag-RDTs are generally reported as being easy to use and low cost, with a rapid turnaround time that enables timely identification of cases and subsequent interventions to prevent onward transmission of COVID-19

*What do the new findings imply?:*

- The evidence indicates that Ag-RDTs can be effectively deployed across a broad range of settings when consideration is given to how they are implemented and interpreted
- The development of more detailed, evidence-based testing policies for Ag-RDTs will be important to help countries implement effective testing programmes and make the best use of Ag-RDTs as part of the COVID-19 testing toolkit

## INTRODUCTION

Testing for SARS-CoV-2 remains a critical tool in controlling the COVID-19 pandemic. Testing allows the early identification of cases, enabling rapid isolation of positive cases and linkage to treatment, as well as monitoring of outbreaks and broader epidemiological surveillance. Antigen-detecting rapid diagnostic tests (Ag-RDTs) that can be used at the point of care have the potential to expand timely access to COVID-19 testing, particularly in settings with limited laboratory capacity or in outbreak settings. As Ag-RDT results can be made available in 15–30 minutes, they have applications in a variety of settings including screening ahead of mass events, rapid testing at ports of entry, and for surveillance, particularly in settings without laboratory facilities for molecular tests.

Ag-RDTs are one of two main classes of diagnostic tools for detecting active SARS-CoV-2 infection, the other being nucleic acid amplification tests (NAATs) such as real time reverse-transcription polymerase chain reaction (rRT-PCR)-based assays.^1 2^ Ag-RDTs work by detecting SARS-CoV-2 antigens produced by the replicating virus in respiratory secretions, while NAATs detect viral RNA.^1 2^ The accuracy of Ag-RDTs is dependent on factors such as the viral load in the specimen, the quality of the sample and time from onset of infection.^2^ While the sensitivity of Ag-RDTs is typically lower than NAATs, they have advantages in terms of their simplicity, low cost and rapid results compared with NAATs, which require laboratory facilities and trained technicians.^1 2^ Ag-RDTs are usually most accurate when viral loads are highest, shortly before and in the first week after symptom onset.^2 3^

A number of Ag-RDTs have received country regulatory approvals and are being deployed in a variety of settings for mass or targeted screening of specific groups like healthcare workers, schoolchildren and travellers. Guidance around the use of Ag-RDTs is varied. The World Health Organization (WHO) has issued interim guidance on the use of Ag-RDTs, recommending that Ag-RDTs meeting the minimum performance requirements of ≥80% sensitivity and ≥97% specificity compared with a NAAT reference assay can be used in settings likely to have the most impact on early detection of cases for care and contact tracing and where test results are most likely to be correct. Priority uses include community testing of symptomatic individuals meeting the suspected COVID-19 case definition, to detect and respond to suspected outbreaks of COVID-19, and to screen asymptomatic individuals at high risk of COVID-19, including health workers, contacts of cases and other at-risk individuals. However, information on country-level testing policies for Ag-RDTs collected by FIND, the global alliance for diagnostics, shows that such policies can vary substantially by country.^4^

While a number of studies have characterized the diagnostic accuracy of Ag-RDTs,^3 5-7^ a comprehensive assessment of how Ag-RDTs are being used in the real world and their performance in each setting has not been conducted to date. In this systematic review, we aimed to synthesize the published and preprint literature regarding specific use cases and overall performance of Ag-RDTs for detection of SARS-CoV-2 in specific settings. Our primary objectives were to understand (i) settings where Ag-RDTs have been used for COVID-19 screening and/or surveillance, ii) and what were their performance characteristics across varied settings.

## METHODS

### Search strategy

Three reviewers (AA, JB, EM) searched two electronic databases of published literature (PubMed, EMBASE) and one preprint database (medRxiv). The search string contained two elements: “COVID-19” and “antigen”. Neither patient databases nor the grey literature were searched. No restrictions were applied on year of publication. The search was run three times, on 14 December 2020, 22 February 2021 and 12 April 2021. The protocol for this review was not registered.

### Inclusion criteria

Studies published in English and French were included. Reference lists of screened full-text publications and reviews were reviewed to identify potential publications that were not found in the original search.

There were two types of use cases of interest in this review. The first was mass screening, irrespective of symptoms across a wide variety of settings, e.g. workplace, schools, universities, airports, essential workers; this included healthcare centres for patient admittance or triage, as well as healthcare workers and hospital staff, if testing was not based on clinical or epidemiological suspicion. The second was surveillance, which included two settings: i) serial or repeat testing of well-defined groups, e.g. students in professional medical training programmes, professional sports franchises (players and staff), and ii) random samples of a large population, all without clinical or epidemiological suspicion of SARS-CoV-2 infection.

Eligible populations included individuals of any age and from any geographic region who had undergone testing with a SARS-CoV-2 Ag-RDT as part of a screening strategy that was not limited exclusively to presumptive patients. Studies were both prospective or retrospective in nature and single- or multiple-gated in terms of enrollment. Studies were not required to utilize a reference test, such as RT-PCR, for inclusion.

### Exclusion criteria

Studies where an Ag-RDT was not the index test of interest were excluded, as were modelling studies. Diagnostic accuracy studies (DAS) that did not use an Ag-RDT in the context of mass screening or surveillance were also excluded, for example, DAS looking solely at Ag-RDT performance at a testing site for people referred for COVID-19 testing by healthcare workers. Studies with a population that consisted only of presumptive COVID-19 patients (e.g. those with symptoms indicative of COVID-19 who requiring testing as part of clinical diagnosis) were therefore also excluded, as this did not fit the criterion of mass screening or surveillance. In addition, case series and studies of Ag-RDTs in the preclinical or analytical validation stages were excluded.

### Screening, study selection, and data extraction

All citations captured by the search were compiled, de-duplicated and managed in Covidence.^8^ All preprints and publications were screened for eligibility by two of three reviewers (AA, JB, EM) by title, abstract, and full-text. Disagreements were resolved through discussion between the same three reviewers. A structured Google form was piloted with a subset of studies and used for data extraction, with double data extraction of each study performed by four authors (AA, JB, EM, TU).

Information on the study setting, population, type of testing, brand of test, diagnostic performance and other outcomes were extracted, along with operational findings and author conclusions. A complete list of extracted fields is shown in Supplementary Table 1. Data entries for each study were reviewed and harmonized by four authors (AA, JB, EM, TU).

### Quality assessment

For DAS, the revised Quality Assessment of Diagnostic Accuracy Studies (QUADAS-2) tool^9^ was adapted to fit the scope of the systematic review and used to assess risk of bias in each of four domains (patient selection, index test, reference standard, and flow and timing). For other studies reporting outcomes such as prevalence, test positivity and effectiveness, a tool based on the approach proposed by Munn et al. 2015 for assessing the quality of prevalence studies^10^ was developed to assess the risk of bias. Definitions for the questions included in each tool are provided in Supplementary Tables 2 and 3. Double quality assessment was performed for each study by two of three authors (AA, JB, TU), with discrepancies resolved through discussion between the same three authors.

### Main outcomes of interest and data synthesis

Due to the heterogeneous nature of the inclusion criteria, several possible outcomes could have been reported by the included studies. These included diagnostic accuracy (sensitivity and specificity), test positivity rate, effectiveness (various definitions), impact (various definitions), and prevalence (various definitions). We also included operational findings that studies reported with respect to test utility, feasibility, and acceptability.

Findings were tabulated and summarized descriptively. Due to the heterogeneous nature of the studies and reporting of outcomes, no meta-analyses were performed. Outcomes are presented using the nomenclature of each study. Other assessments of Ag-RDT diagnostic accuracy exist and we invite readers to consult them regarding the performance of Ag-RDT in various settings.^3 5-7^

### Patient and public involvement

Patients and the public were not involved in this study.

## RESULTS

After deduplication, 4313 studies from 2020 to 2021 were included (Figure 1). A total of 4147 studies were excluded after title/abstract screening and 165 full-text publications were assessed. Ultimately, 39 studies were included in our qualitative synthesis. Publications were most commonly excluded because they were DAS that did not use Ag-RDTs in the context of mass screening or surveillance, reviews or modelling studies (Figure 1).

**Figure 1.**
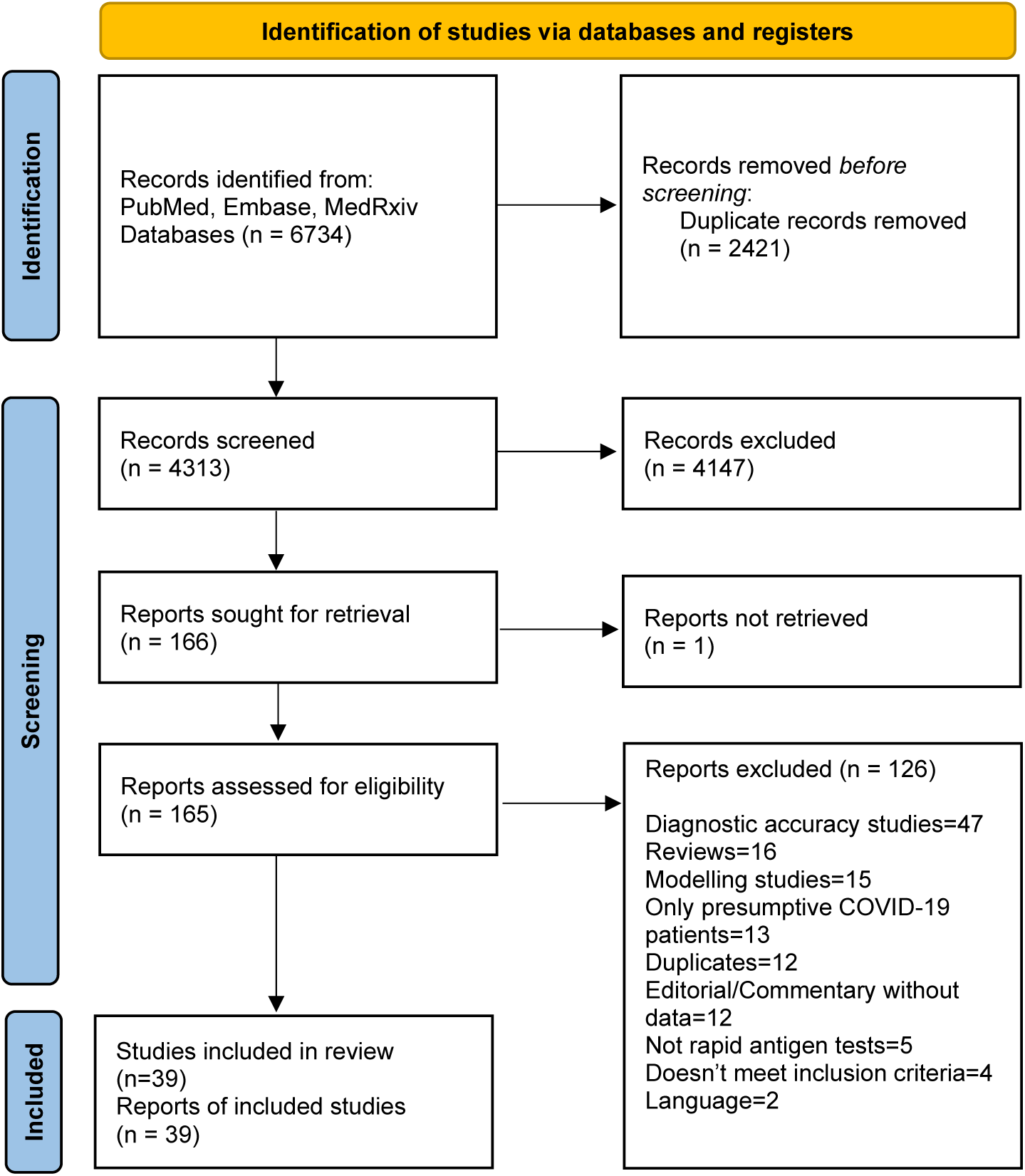
PRISMA study flowchart.

Of the final subset of 39 studies, 35 were published and four were preprints at the time the review was conducted. A majority of studies were conducted in high-income countries, with 13/39 studies from the USA. Four of the 39 papers were conducted in low- and middle-income countries (LMICs), with three studies in India and one in Cameroon.

A single-gate design – where all participants were recruited according to a single set of criteria^11^– was used in 35 of 39 studies; three studies utilized a phased intervention, and one study was two-gated. Different sampling strategies were employed in the studies, the most common being consecutive sampling (utilized in 12/39 studies). However, 18 studies did not describe their sampling scheme in a sufficiently clear manner to allow judgement.

### Quality assessment

Quality assessments for the studies that reported various outcomes (diagnostic performance and others) are presented in Tables 1 and 2, respectively.

**Table 1.** Quality assessment for diagnostic accuracy studies using modified QUADAS-2.

| Study (First author) | Risk of bias |  |  |  |  |  |  |  | Applicability |  |
| --- | --- | --- | --- | --- | --- | --- | --- | --- | --- | --- |
|  | Was a consecutive or random sample of patients enrolled? | Was a case-control design avoided? | Did the study avoid inappropriate exclusions? | Is the reference standard likely to correctly classify the target condition? | Was there an appropriate interval between index tests and reference standard? | Did all patients receive a reference standard? | Did all patients receive the same reference standard? | Were all patients included in the analysis? | Are there concerns that the index test, its conduct, or its interpretation differ from the review question? | Are there concerns that the target condition as defined by the reference standard does not match the review question? |
| Alemaný<sup>56</sup> | ? | + | ? | + | ? | + | + | + | + | + |
| Turcato<sup>32</sup> | + | + | + | + | + | + | + | + | + | + |
| Igloi<sup>17</sup> | + | + | + | + | + | + | + | + | + | + |
| Van Honacker<sup>33</sup> | + | + | + | + | + | + | + | + | + | + |
| Gili<sup>13</sup> | ? | + | + | + | + | + | + | + | + | + |
| Seitz<sup>45</sup> | ? | + | ? | + | + | + | + | – | + | + |
| Sood<sup>46</sup> | ? | + | + | + | + | + | + | + | + | + |
| Pilarowski<sup>20</sup> | ? | + | ? | + | + | + | + | + | + | + |
| Smith<sup>41</sup> | ? | – | + | + | + | + | + | – | + | + |
| Pollock<sup>18</sup> | + | + | + | + | + | + | + | + | + | + |
| Pollock<sup>19</sup> | + | + | + | + | + | + | + | + | + | + |
| Boum<sup>29</sup> | + | + | + | + | ? | + | + | + | + | + |
| Pilarowski<sup>21</sup> | – | + | + | + | + | + | + | + | + | + |
| Okoye<sup>24</sup> | ? | + | + | + | + | + | + | + | + | + |
| Pray<sup>25</sup> | ? | + | + | + | + | + | + | + | + | + |
| Shah<sup>22</sup> | ? | + | + | + | + | + | + | + | + | + |
| James<sup>35</sup> | + | + | + | + | + | + | + | + | + | + |
| Rottenstreich<sup>30</sup> | + | + | + | + | + | + | + | + | + | + |
| Regev-Yochay<sup>14</sup> | ? | + | + | + | + | + | + | + | + | + |
| Winkel<sup>44</sup> | ? | + | + | + | ? | – | + | + | + | + |
| Kriemler<sup>43</sup> | – | + | – | + | + | + | + | + | + | + |
| Cerutti<sup>23</sup> | ? | + | ? | + | + | + | + | + | + | + |

Low risk Unclear risk High risk

**Table 2.**
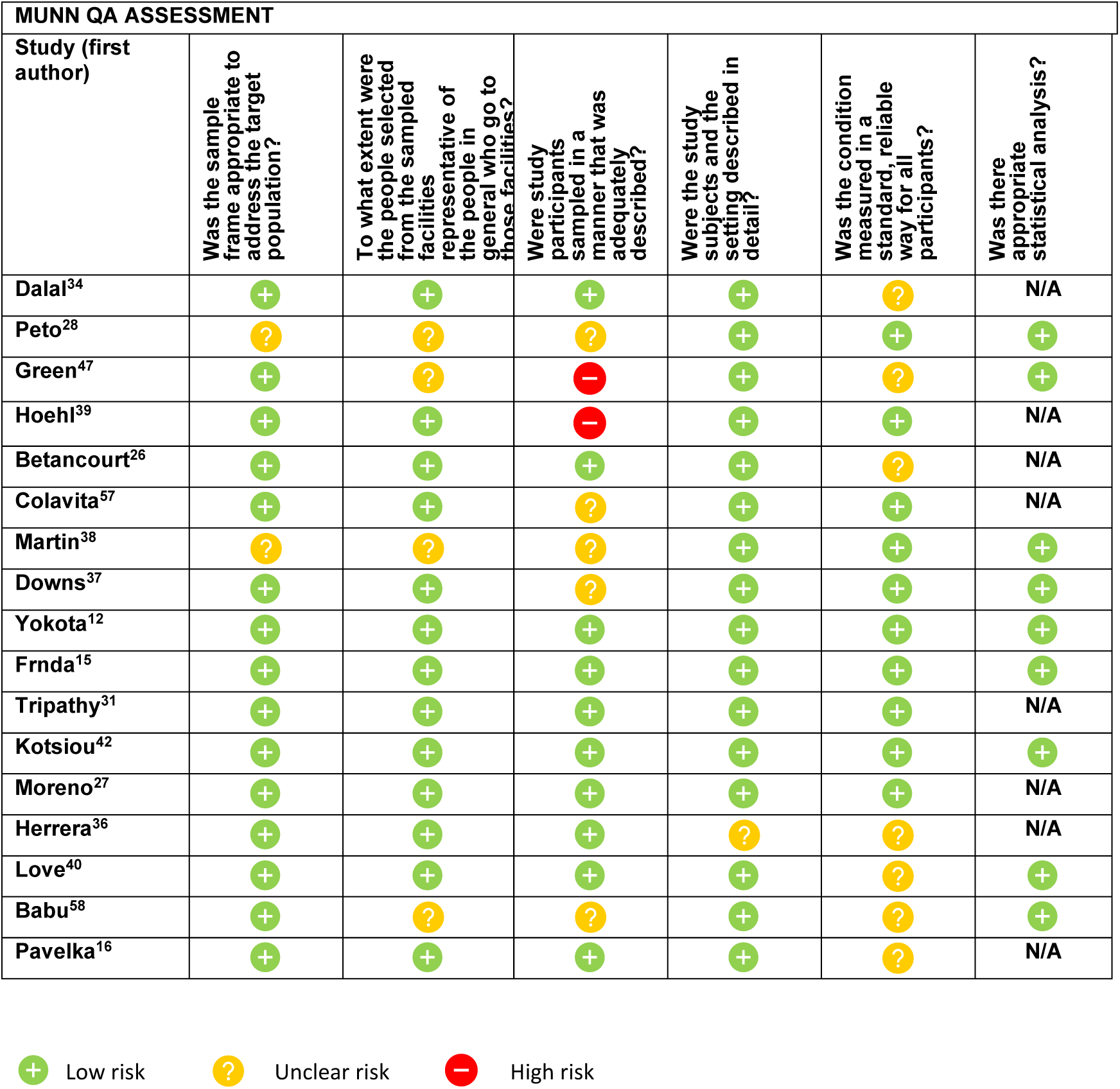
Quality assessment for non-diagnostic accuracy studies adapted from Munn et al.

For diagnostic accuracy studies, two of the 22 studies were assessed as having a high risk of bias due to sampling, and 12 out of the 22 (54.5%) studies had an unclear risk of bias due to sampling, as the specific sampling schemes used in the studies were not explicitly described in the paper (Table 1). Risk of bias was generally low across other aspects of the patient selection domain, and across the index test, reference standard, and flow and timing domains. All studies had low applicability concerns (Table 1).

For studies reporting on other outcomes, risk of bias was also generally low or unclear. Risk of bias was high for two studies in relation to the sampling scheme, as it was not adequately described in the respective articles. Risk of bias was unclear in 7/17 studies (41.2%) in relation to whether the condition was measured in a standard, reliable way for all participants, as it was not clear if the Ag-RDT had been performed using the same procedure for all participants.

### Results of individual studies

Table 3 shows the results from individual studies. All studies but two investigated Ag-RDTs in the standalone strip format (lateral flow); two studies investigated the use of a multiplex test (sandwich chemiluminescent enzyme immunoassay).^12 13^ The brand of test was reported for 36 of the 39 studies and two studies investigated the use of multiple brands of Ag-RDTs.^14 15^

**Table 3.**
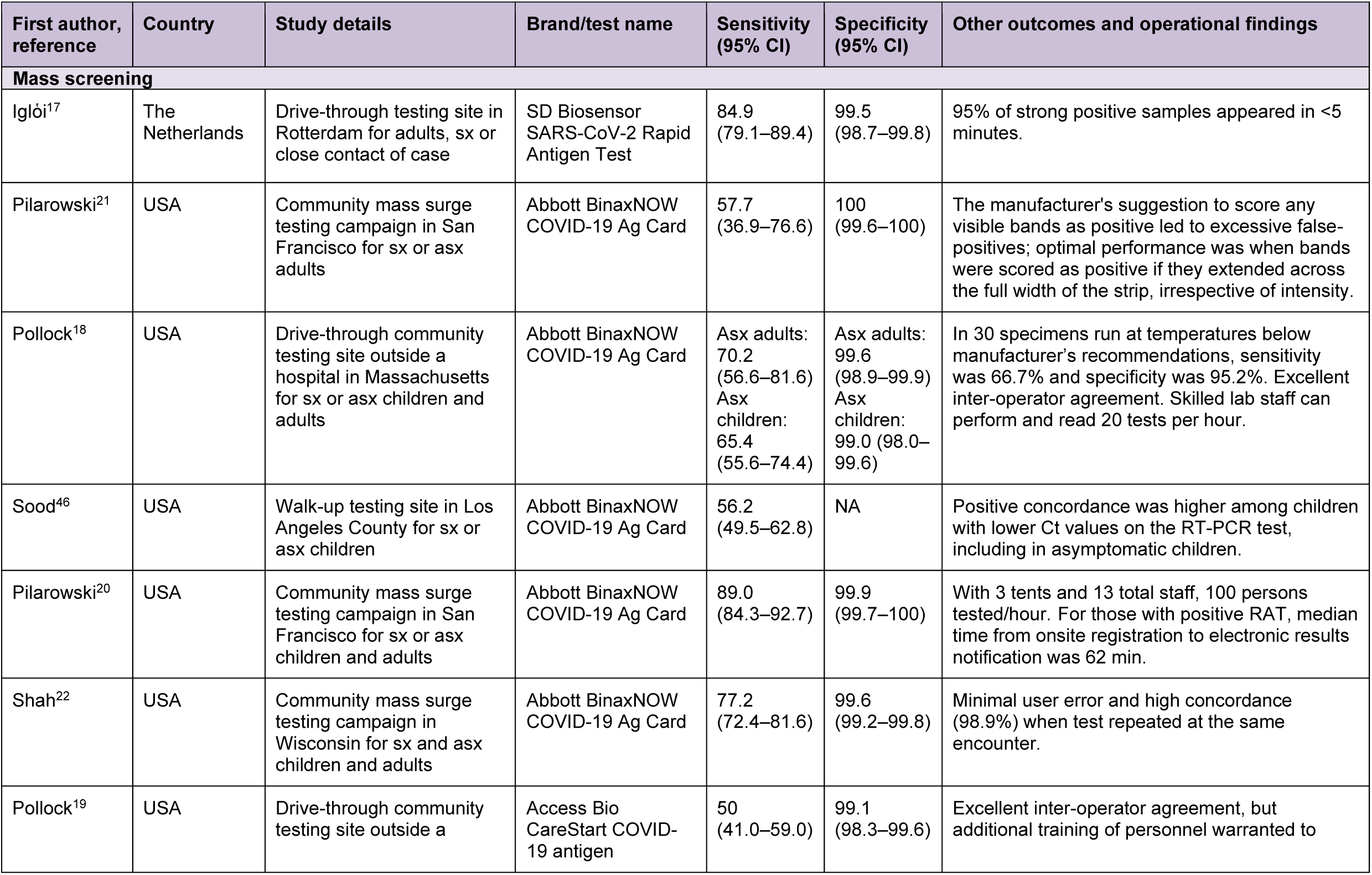
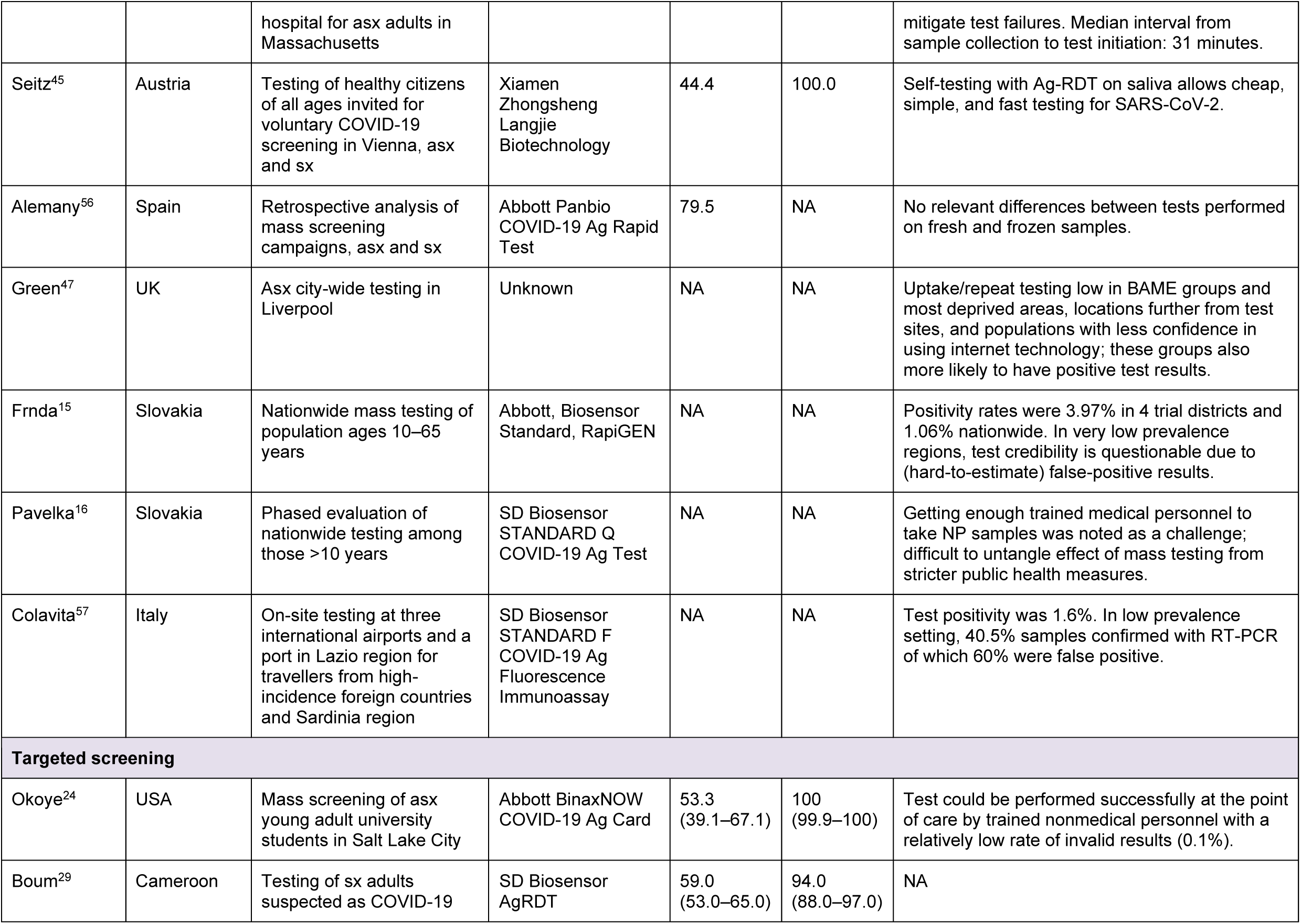
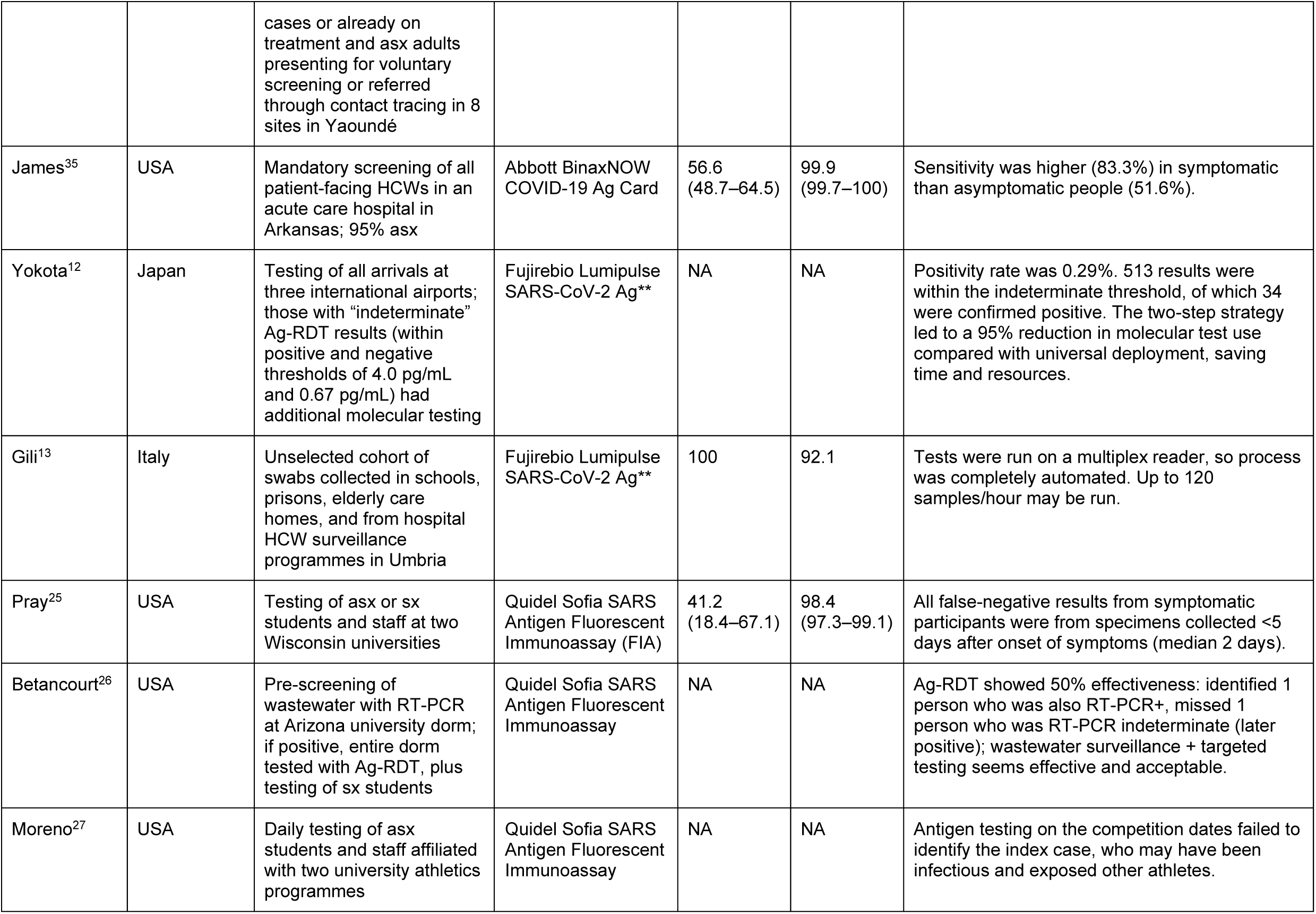
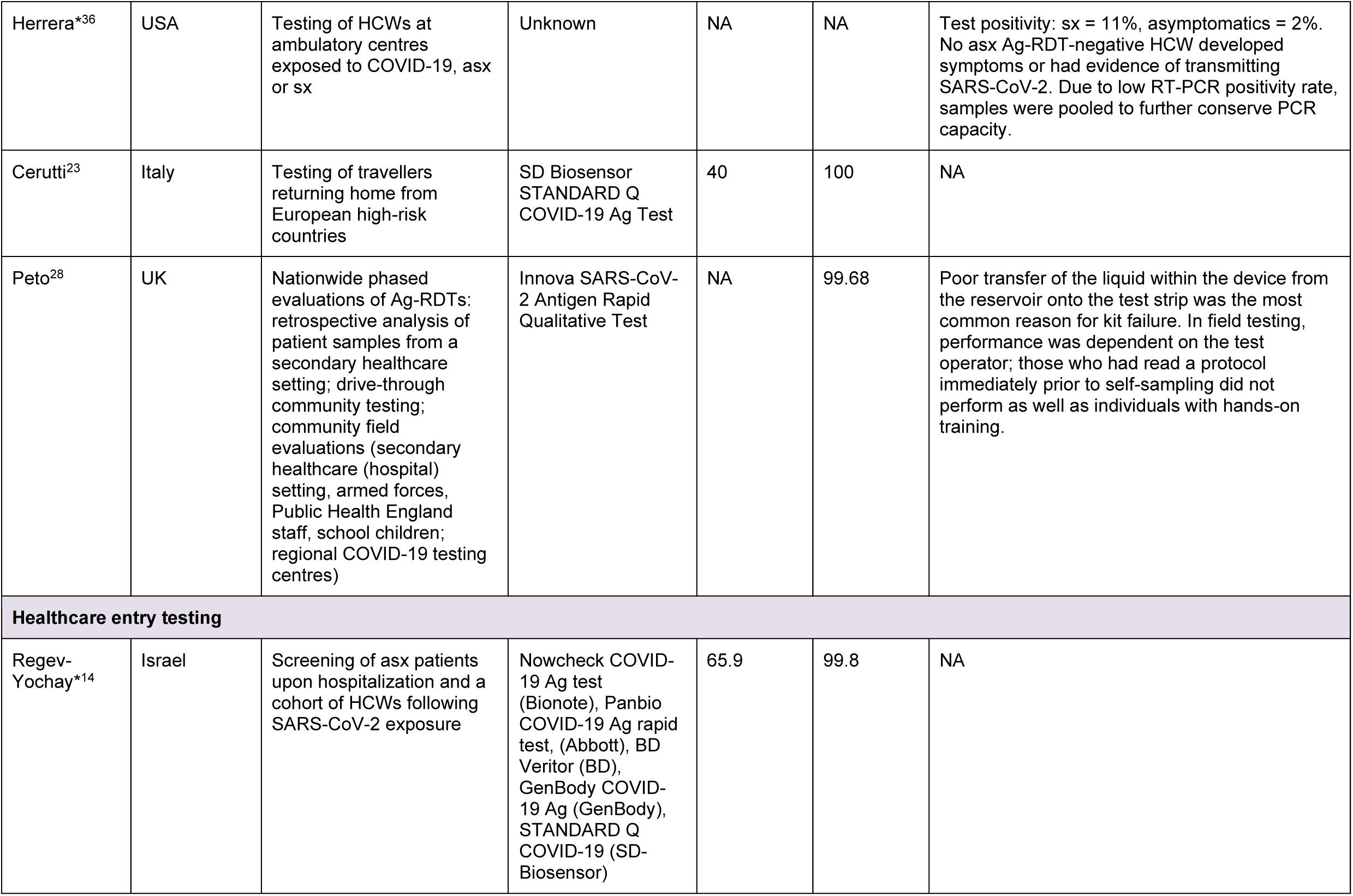
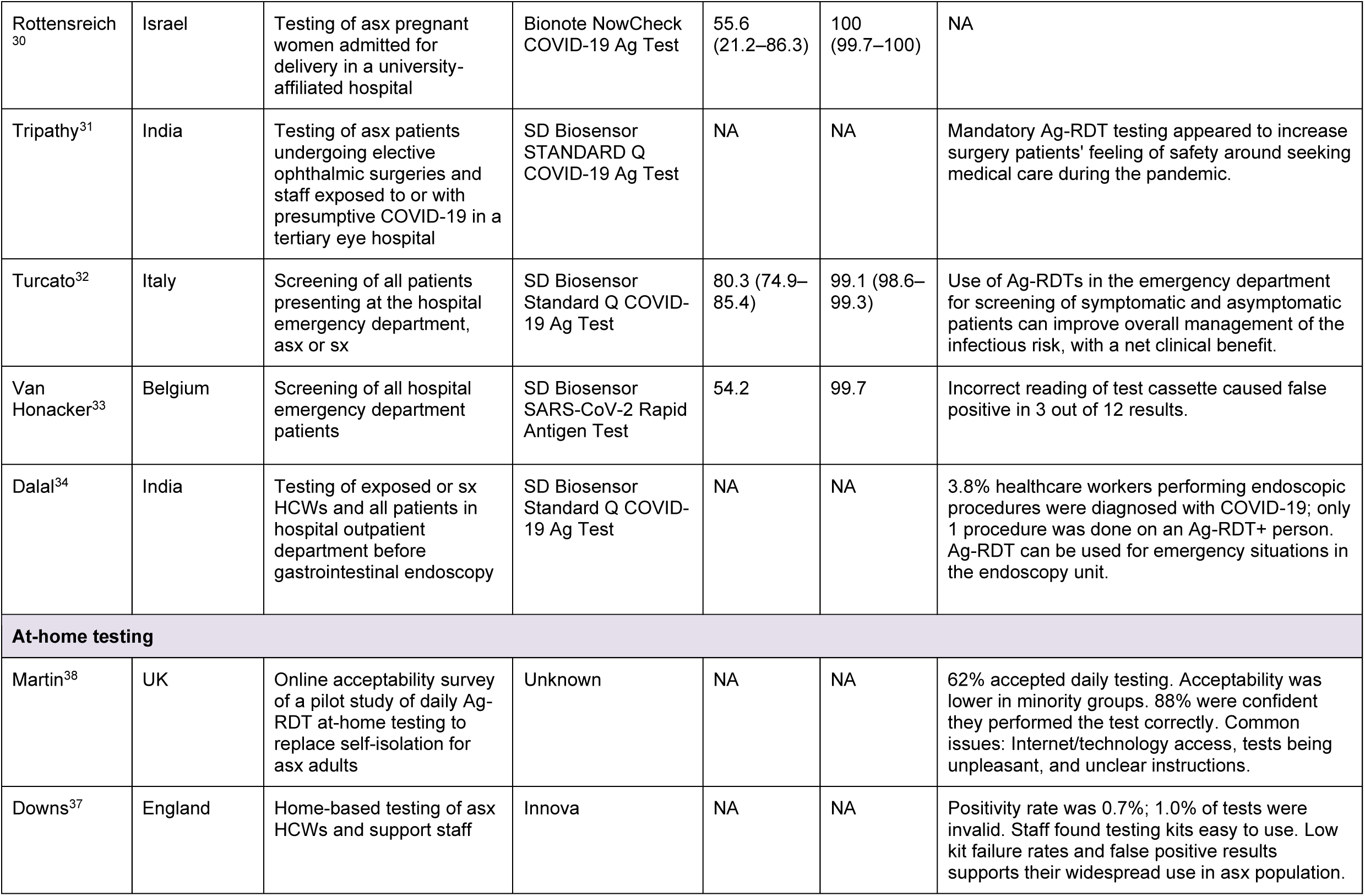
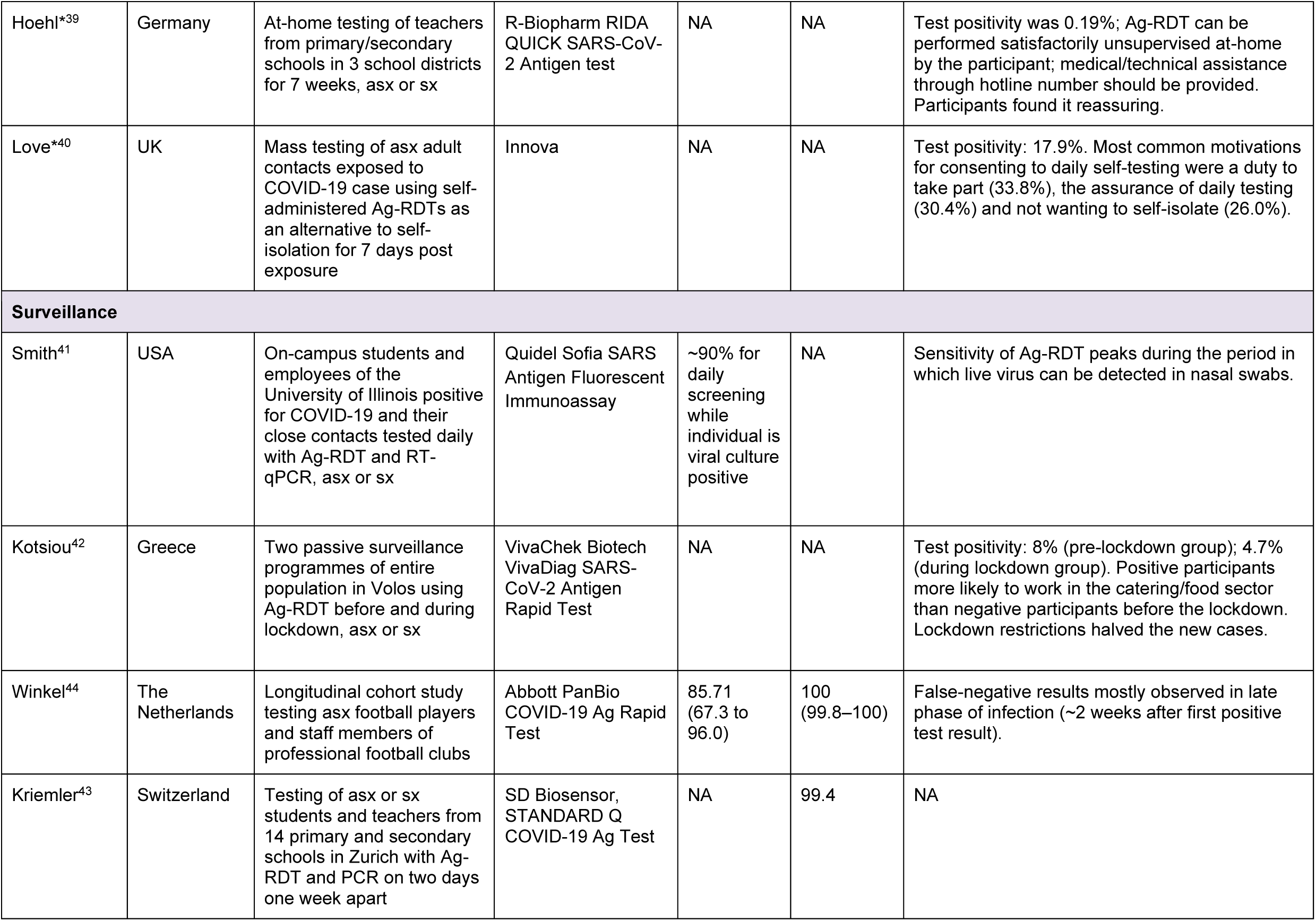
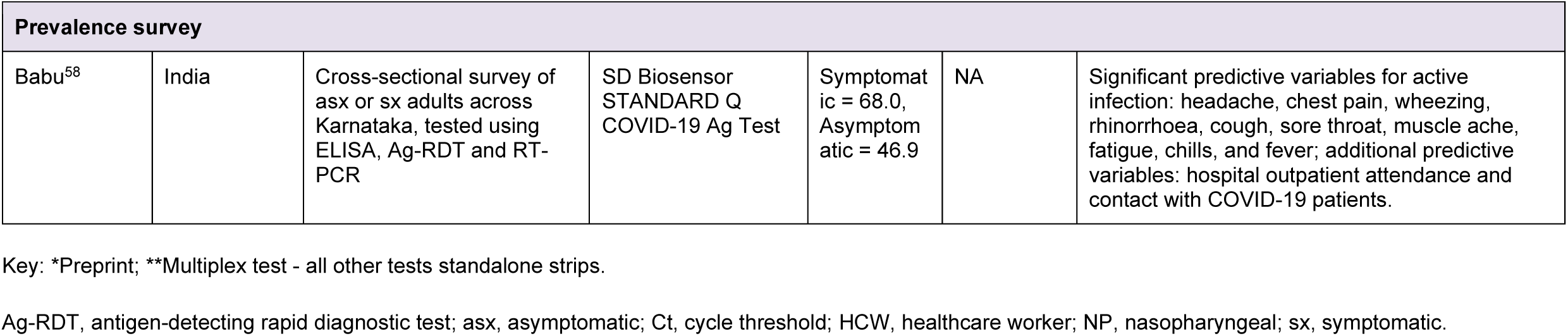
Individual study results: Ag-RDT use cases, performance and operational findings.

#### Type of testing

Six broad types of screening initiatives were identified: mass screening, targeted screening, healthcare entry testing, at-home testing, surveillance and prevalence surveys. Figure 2 shows the range of study sizes across the different testing types.

**Figure 2.**
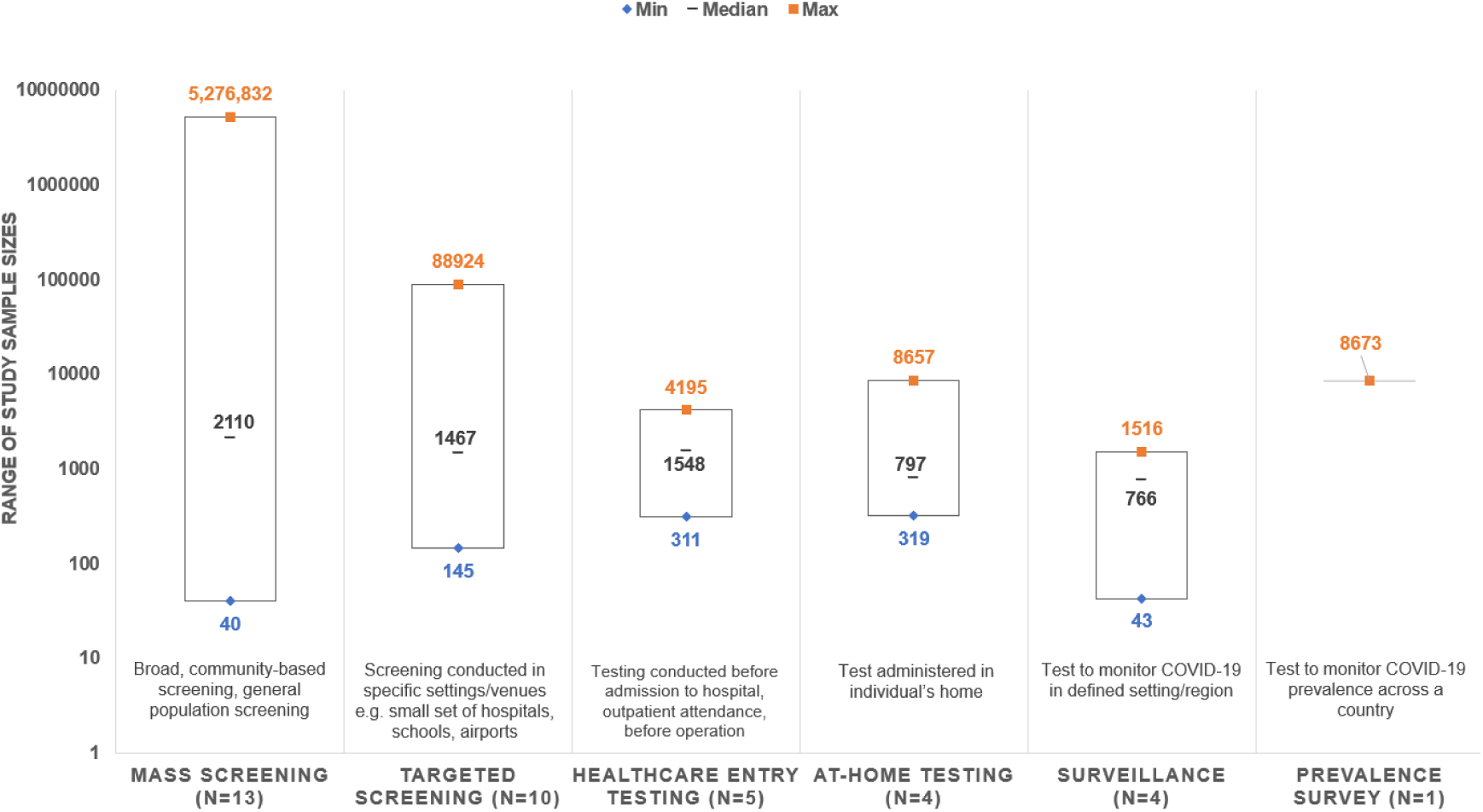
Range of sample sizes across each type of testing. Note: Sample size was not reported for one study of targeted screening and one study of healthcare entry testing.

Mass screening was used to describe non-targeted testing interventions such as broad community-based screening and population screening. Targeted screening was used to describe screening conducted in specific settings/venues, such as a small set of hospitals, schools, or airports. Testing administered in an individual’s home was described as at-home testing, while testing before admission to/upon attendance at a hospital was described as healthcare entry testing. Studies to monitor COVID-19 in a defined setting or region were referred to as surveillance studies, while those looking at COVID-19 prevalence across a country were referred to as prevalence surveys.

The most common use of Ag-RDTs, in 13 studies, was for mass screening interventions, which included nationwide and citywide screening efforts, community hotspot testing and testing of travellers. Specific types of mass screening included two studies reporting multiple rounds of nationwide testing in Slovakia, where millions of individuals – over 80% of the age-eligible population – were tested using Ag-RDTs.^15 16^ Mass screening efforts also included three studies of drive-through testing in the USA and the Netherlands^17-19^, and three studies of community mass surge testing in the USA^20-22^ among others.

The second most common type of testing was targeted screening of specific populations, including testing at airports and ports of entry,^12 23^ universities (campuses, dormitories, athletics programmes) ^24-27^, and mixed interventions.^13 28 29^

Six studies described testing interventions aimed at individuals seeking healthcare, including ahead of emergency room admittance, outpatient attendance, baby deliveries, and surgery.^14 30-34^ Testing efforts specifically targeted at healthcare workers were described in five studies.^14 34-37^

Four studies described self-testing efforts that were carried out at-home; study populations included schoolteachers and hospital staff in two studies, and asymptomatic adult contacts of COVID-19 cases in the UK who were offered home tests in the other two studies.^37-40^ For home-testing studies, tests were given to participants with instructions for use and participants were asked to self-report their test results.

Four surveillance studies were conducted in Greece, Switzerland, USA, and the Netherlands, which used Ag-RDTs to monitor COVID-19 across an entire town, primary and secondary schoolchildren and teachers, university students and staff, and in professional football clubs, respectively.^41-44^

#### Sample type

Sample type was reported in all but six studies (Supplementary Table 4). Nasal or nasopharyngeal swabs were used in a vast majority of studies that reported sample type, either alone or in combination with another sample type (30/33 studies). One study investigated the use of buccal swabs in schoolchildren in Switzerland.^43^ Two studies investigated the use of saliva samples: the first as part of a mass screening effort in Austria,^45^ and the other for testing arrivals at three international airports in Japan.^12^

#### Sensitivity of Ag-RDTs

Across studies that reported diagnostic accuracy, sensitivity of Ag-RDTs varied:

a. Mass screening from 44.4% to 89.0% (n=9)
b. Targeted screening from 40% to 100% (n=6)
c. Healthcare entry testing from 54.2% to 80.3% (n=4)
d. Surveillance from 85.7% to ∼90% (n=2)
e. In the single prevalence survey, sensitivity was 68.0% among symptomatic individuals and 46.9% among asymptomatic individuals
f. None of the studies of at-home testing provided estimates of sensitivity.

A trend for higher sensitivity among symptomatic versus asymptomatic individuals tested with Ag-RDTs was frequently observed, with lower sensitivity during early asymptomatic infections and in the later phase of infection.

Consequently, some studies recommended confirmatory testing with PCR when using Ag-RDTs.^12 25 35 45^ Studies where participants also underwent testing with PCR generally reported that Ag-RDTs performed better in individuals with relatively lower PCR cycle threshold values, indicating higher viral load, compared with groups in the same study with higher cycle threshold values.^18 19 46^

Serial testing with Ag-RDTs was noted as a potential strategy to help compensate for limited sensitivity during early infection in some^37 46^ but not all^27^ of the studies that investigated this approach.

### Operational findings

Overall, Ag-RDTs were generally noted as easy to use and low cost, with a rapid turnaround time that enabled timely identification of cases and subsequent interventions to prevent onward transmission of COVID-19. Ag-RDTs were noted as a useful tool in large-scale screening of mixed populations (asymptomatic and symptomatic) and to screen asymptomatic individuals, particularly in high-prevalence settings, and in settings with limited resources.

In terms of the interpretation of the Ag-RDT result, various studies noted minimal user errors when the tests were conducted by trained personnel/healthcare workers or by participants themselves. However, studies emphasized the importance of training and clear instruction, with one study finding that field performance depended on the operator: the Ag-RDTs performed better in those with hands-on training than in those without.^28^

Another study identified that temperature/climate conditions can affect Ag-RDT performance, highlighting the importance of conducting Ag-RDTs in controlled conditions.^18^ Test-specific findings were also reported, including that “strong” positive bands tended to appear quickly^17^ and that optimal performance was achieved when scoring bands as positive on the lateral flow format Ag-RDTs when they extended across the full width of the strip, irrespective of intensity (rather than the manufacturer’s recommendation to score any visible band as positive).^21^

The studies evaluating at-home testing interventions noted generally high acceptability and compliance with self-testing.^37-40^ Reports found that self-testing was usually performed to a satisfactory standard and that the majority of participants felt confident that they performed the test correctly.^37-40^ In one study of at-home testing among schoolteachers, participants reported that regular testing with Ag-RDTs was reassuring when working in schools during the pandemic.^39^ However, inequalities in the uptake of Ag-RDTs were identified, with lower uptake in deprived areas, ethnic minorities, areas with limited access to test sites and those with low digital access.^47^

## DISCUSSION

The availability of SARS-CoV-2 Ag-RDTs has substantially broadened testing approaches for COVID-19, particularly in decentralized settings. This systematic review presents the first detailed assessment of Ag-RDTs use cases, in terms of how Ag-RDTs have been used for COVID-19 screening and surveillance since the start of the pandemic. Our findings demonstrate that Ag-RDTs are being used across a wide range of real-world settings, including as part of mass and targeted screening efforts, healthcare entry testing, at-home testing, surveillance and prevalence studies.

Overall, the studies included in this assessment have reported Ag-RDTs to be convenient, rapid, and low-cost interventions that can increase access to testing in a variety of settings to respond to changing testing needs. Importantly, the review identified that Ag-RDTs could be performed and interpreted correctly when conducted by healthcare workers, other trained personnel and by participants themselves across the different use cases.

The sensitivity of Ag-RDTs across studies in the review was variable compared with RT-PCR, and in some cases low, particularly among asymptomatic individuals. Sensitivity estimates ranged from 40% to 100% across the studies, which included either asymptomatic or mixed (asymptomatic and symptomatic) populations. Despite the lower sensitivity of Ag-RDTs compared with RT-PCR, a number of the studies noted the value of testing with Ag-RDTs, particularly in high-prevalence settings and where testing resources are limited, as a means to identify infected individuals that would otherwise be missed. However, certain studies did advise caution when using Ag-RDTs in situations where high test sensitivity is critical, for example in hospital emergency departments, particularly without confirmatory testing.^33 44^

The lower sensitivity of Ag-RDTs in asymptomatic individuals aligns with findings from meta-analyses of Ag-RDT diagnostic accuracy. In a Cochrane review, commercially available Ag-RDTs correctly identified SARS-CoV-2 infection in an average of 72% of people with symptoms, compared with 58% of people without symptoms.^3^ Another systematic review found similar results, with a pooled sensitivity of 76.7% in symptomatic patients versus 52.5% in asymptomatic patients.^7^

While this review demonstrates the diverse settings in which Ag-RDTs have been used, the variable performance of Ag-RDTs compared with RT-PCR means that the value of testing with this approach needs to be carefully evaluated for each use case. The prevalence of COVID-19 in a population is a particularly important consideration, as it affects the predictive value of the test; for example, the probability that a positive Ag-RDT result is a true positive is reduced in low prevalence settings. This has implications for what can be a delicate cost-benefit balance in terms of the societal benefits of the tests. Each false positive result in the real world may result in an entire classroom or a large part of workplace having to isolate at home for 10–14 days. In a healthcare setting, a high false positive rate may put undue pressure on health systems through the number of staff having to isolate.^48^ In contrast, false negative results may enable a SARS-CoV-2-infected person to get on an airplane or attend a mass event.

Nevertheless, it has been pointed out that these costs should be considered in terms of the broader picture, where without strategies to break chains of COVID-19 transmission on a smaller scale – in workplaces, in schools – governments have had to resort to lockdowns of entire cities and countries that have come at an enormous cost to individuals, societies and economies.^49^ Consequently, careful evaluation of each situation is needed to determine how Ag-RDTs can be optimally implemented to improve COVID-19 control. Important factors to consider include the prevalence of COVID-19 in the population, the consequences of false positive or false negative results, and how testing should be applied as part of testing algorithms (e.g. whether serial testing can mitigate low sensitivity, whether confirmatory testing of positive or negative Ag-RDT results with RT-PCR is required). WHO interim guidance notes that Ag-RDTs are most reliable in areas where there is ongoing community transmission of COVID-19 (≥5% test positivity rate) and advise the use of NAATs for first-line testing or confirmation of positive Ag-RDT results in low transmission settings.^1^

When considering the overall value of Ag-RDTs for large-scale screening and surveillance, consideration should also be given to whether the sensitivity of Ag-RDTs is of secondary importance to other characteristics such as turnaround time and accessibility. Indeed, a study modelling the effectiveness of repeated population screening for SARS-CoV-2 found that effective screening depended largely on the frequency and speed of testing and was only marginally improved by high test sensitivity.^50^ In addition, there is evidence from the USA that Ag-RDTs have a higher positive predictive value than RT-PCR for active SARS-CoV-2 infection,^51^ despite their generally lower overall sensitivity. This is important when evaluating the suitability of tests for widespread screening, where the priority is to quickly detect the most infectious individuals to prevent onwards transmission. In contrast, RT-PCR can detect viral RNA for prolonged periods beyond when live virus can be cultured – meaning that people can continue to test positive for SARS-CoV-2 by RT-PCR after they are no longer infectious.^52^

In terms of implications for testing policies, the results of this study demonstrate that there is demand for and use of Ag-RDTs as practical tools to detect SARS-CoV-2 across a broad range of settings and use cases. In particular, Ag-RDTs are being used for widespread screening of asymptomatic individuals beyond the scenarios outlined in interim global guidance from WHO. These findings align with the broader picture of COVID-19 testing emerging over the last year, which has seen calls for wider use of Ag-RDTs from public health experts^49 52 53^ and the implementation of expansive programmes with Ag-RDTs in high-income countries like the UK and Canada for testing of the general population.^53-55^

It’s clear that Ag-RDTs will have an important role to play in COVID-19 testing moving forward, but more detailed, evidence-based testing policies around the use of Ag-RDTs will be critical to help countries implement effective testing programmes for their settings. The findings from this work along with additional evidence generation, particularly in LMICs, can help inform more comprehensive and standardized policies.

Considering the systematic review as a whole, strengths include its comprehensive approach and inclusion of a broad range of studies from countries across Europe, North America, Africa and Asia. The review also included a number of studies with large sample sizes (21 of which had a sample size >1000). The systematic review was also conducted using rigorous methods, with study selection, data extraction and quality assessment performed and validated by multiple authors.

Some limitations of the approach are that the studies identified from the review are primarily from the USA and Europe, with fewer reports from LMICs. This limits understanding of how Ag-RDTs have been used across LMICs, but may also reflect that Ag-RDTs have been less available for widespread use in LMICs than in high-income countries.

Heterogeneity in study design and setting also prevented us from undertaking a meta-analysis and limits the extent to which individual study results can be compared. The search was also restricted to English and French, and did not include the grey literature, which may have excluded certain reports. Nevertheless, the review represents the first comprehensive assessment of how Ag-RDTs have been used in the real world.

## CONCLUSIONS

This first detailed assessment of use cases for Ag-RDTs demonstrates their application across a wide range of real-world settings, including as part of mass and targeted screenings, healthcare entry testing, at-home testing, surveillance and prevalence studies. The approaches captured in this review highlight the versatility of Ag-RDTs as rapid, low cost, and easy to use COVID-19 screening tools for asymptomatic and symptomatic individuals. Although the sensitivity of Ag-RDTs can be low compared with RT-PCR, evidence indicates that Ag-RDTs can be easily deployed in a broad range of settings when consideration is given to how they are implemented and interpreted. The development of more detailed, evidence-based testing policies around the use of Ag-RDTs will be important to help countries implement effective testing programmes to help make the best use of Ag-RDTs as part of the COVID-19 testing toolkit.

## Data Availability

The datasets used and/or analysed during the current study are available from the corresponding author on reasonable request.

## DECLARATIONS

### Contributors

The study was conceived by AA, JB, SGS, EM, and AT. The systematic review was performed by AA, JB, EM, and TU as detailed in the methods section. The manuscript drafts were developed by AA, EM, JB, and TU with input from the other authors. All authors contributed to interpretation of data and editing of the article and approved the final version of the manuscript.

### Funding

Funding for the study was provided by FIND; however, ultimate responsibility for opinions and conclusions in the article lies with the authors.

### Competing interests

AT and SC are employed by FIND and SGS was employed by FIND at the time of the study. TU is a consultant for FIND. AA and NPP report a grant from the Canadian Institutes of Health Research for a project on COVID-19 testing unrelated to this study and copyright for a digital program for COVID-19 self-testing. FIND is a not-for-profit foundation that supports the evaluation of publicly prioritized tuberculosis assays and the implementation of WHO-approved (guidance and prequalification) assays using donor grants. FIND has product evaluation agreements with several private sector companies that design diagnostics for tuberculosis and other diseases. These agreements strictly define FIND’s independence and neutrality with regard to these private sector companies.

