## Supplementary Table 1 for "Use cases for COVID-19 screening and surveillance with rapid antigen-detecting tests: a systematic review"

### SUPPLEMENTARY MATERIALS

#### Supplementary Table 1. List of information extracted for each publication

| | |
|---|---|
| **Study identifiers** | • Publication status (preprint or published)<br>• Title of paper<br>• Year of publication<br>• First author<br>• DOI |
| **Study setting** | • Country of study<br>• City or region of study<br>• Type of study setting as described in paper |
| **Study recruitment and sampling scheme** | • Study recruitment scheme<br>• Notes on study recruitment scheme<br>• Sampling scheme<br>• Notes on sampling scheme |
| **Type of testing effort** | • Type of screening/testing effort<br>• Was there repeated/longitudinal testing of the same population? |
| **Study population** | • Was the population a "resident" population (i.e. patients in a care home, prisoners)?<br>• Type of study population<br>• Notes on study population<br>• Age groups<br>• Symptom status<br>• Duration of symptoms (days)<br>• Number of samples considered |
| **Tests** | • Brand of Ag-RDT<br>• Name of Ag-RDT<br>• Type of Ag-RDT used (as stated)<br>• Sample used<br>• Who took the sample<br>• Who ran the Ag-RDT<br>• Was the Ag-RDT reportedly performed according to manufacturer's instructions?<br>• Notes on how Ag-RDT was performed<br>• Were control samples taken before the pandemic?<br>• Were samples used fresh or frozen?<br>• What was the reference standard or comparator test (if any)?<br>• Notes on reference standard/comparator test |

| | |
|---|---|
| **Quantitative results: diagnostic accuracy and other outcomes** | • Number of index test positive<br>• Number of index test negative<br>• Number of reference standard/comparator test positive<br>• Number of reference standard/comparator test negative<br>• Number of indeterminate results<br>• Number of invalid results<br>• Type of measure of effect<br>• Notes on measure of effect<br>• Sensitivity<br>• 95% CI for sensitivity<br>• Specificity<br>• 95% CI for specificity<br>• Impact of Ct value on diagnostic accuracy<br>• Impact of time since symptom onset on diagnostic accuracy<br>• Notes on diagnostic accuracy<br>• Effectiveness/impact<br>• 95% CI for effectiveness/impact<br>• Notes on effectiveness/impact<br>• Prevalence<br>• 95% CI for prevalence<br>• Notes on prevalence<br>• Probability of being infectious<br>• 95% CI for probability of being infectious<br>• Notes on probability of being infectious<br>• Test positivity (not sensitivity)<br>• 95% CI for test probability<br>• Notes on test probability<br>• Other measure of effect<br>• 95% CI for other measure of effect<br>• Notes on other measure of effect |
| **Other** | • Operational findings<br>• Author conclusions<br>• Data extractor comments |

Ag-RDT, antigen-detecting rapid diagnostic test; CI, confidence interval; Ct, cycle threshold.

**Supplementary Table 2. Quality assessment for diagnostic accuracy studies using modified QUADAS-2**

| Question | Definition |
|---|---|
| **Patient selection domain** | |
| RISK OF BIAS: Was a consecutive or random sample of patients enrolled? | • **Yes:** if the paper explicitly stated that a consecutive or random sample of patients was enrolled, or if this was clear from the description of the enrollment<br>• **No:** if the study used a different sampling method (e.g. convenience, purposive)<br>• **Unclear:** if sampling scheme not reported or unclear |
| RISK OF BIAS: Was a case–control design avoided? | • **Yes:** if the study avoided a case–control design<br>• **No:** if the study employed a case–control design<br>• **Unclear:** if study design not reported or unclear |
| RISK OF BIAS: Did the study avoid inappropriate exclusions? | • **Yes:** if no exclusions judged to be inappropriate were reported<br>• **No:** if inappropriate exclusions were noted; for example, if results were only presented for a sub-set of enrolled patients without rationale<br>• **Unclear:** if unclear whether the study avoided inappropriate exclusions |
| **Index test domain** | |
| APPLICABILITY: Are there concerns that the index test, its conduct, or its interpretation differ from the review question? | • **Yes:** if the Ag-RDT was conducted or interpreted for something other than detection of current SARS-CoV-2 infection<br>• **No:** if the Ag-RDT was performed to detect current SARS-CoV-2 infection<br>• **Unclear:** if the purpose of the conduct or interpretation of the Ag-RDT was unclear |
| **Reference standard domain** | |
| RISK OF BIAS: Is the reference standard likely to correctly classify the target condition? | • **Yes:** if the reference standard is suitable for detection of current SARS-CoV-2 infection e.g. PCR-based reference standard or viral culture<br>• **No:** if the reference standard is not suitable for detection of current SARS-CoV-2 infection<br>• **Unclear:** if the reference standard is not described |
| APPLICABILITY: Are there concerns that the target condition as defined by the reference standard does not match the review question? | • **Yes:** if target condition is not SARS-COV-2/COVID-19<br>• **No:** if the target condition is SARS-CoV-2/COVID-19<br>• **Unclear:** if the target condition is not described |
| **Flow and timing domain** | |
| RISK OF BIAS: Was there an appropriate interval between index tests and reference standard? | • **Yes:** if the interval between sample collection for Ag-RDT and reference standard was deemed to be appropriate (e.g. collection at the same visit or within ~1 day of sample for Ag-RDT)<br>• **No:** If the interval between sample collection for Ag-RDT and reference standard was deemed to be inappropriate (e.g. collection of sample for PCR a week after Ag-RDT) |

| | |
|---|---|
| | • **Unclear:** if interval between Ag-RDT and reference standard not reported or unclear |
| RISK OF BIAS: Did all patients receive a reference standard? | • **Yes:** if all patients received reference standard for SARS-CoV-2 infection e.g. PCR or viral culture<br>• **No:** if all patients did not receive a reference standard for SARS-CoV-2 infection<br>• **Unclear:** if unclear whether all patients received a reference standard |
| RISK OF BIAS: Did all patients receive the same reference standard? | • **Yes:** if all patients received the same reference standard for SARS-CoV-2 infection e.g. PCR (note: PCR could be performed in different laboratories)<br>• **No:** if patients received different reference standards (e.g. some received PCR testing, some received viral culture only)<br>• **Unclear:** if unclear whether all patients received the same reference standard |
| RISK OF BIAS: Were all patients included in the analysis? | • **Yes:** if all patients were included in the final analysis<br>• **No:** if some patients were excluded from the final analysis<br>• **Unclear:** if unclear whether all patients were included in the analysis |

**Grading for risk of bias questions:** Yes = low risk; No = high risk; Unclear = unclear risk.

**Grading for applicability questions:** Yes = high risk; No = low risk; Unclear = unclear risk; NA: question not applicable.

**Supplementary Table 3. Definitions for quality assessment for non-diagnostic accuracy studies adapted from Munn et al.**

| Question | Definition |
|---|---|
| Was the sample frame appropriate to address the target population? | • **Yes:** if the sampled facilities are representative of the typical facilities in the area<br>• **No:** if the sampled facilities are not representative of the typical facilities in the area<br>• Unclear: if unclear whether the sampled facilities are representative of those in the area |
| To what extent were the people selected from the sampled facilities representative of the people in general who go to those facilities? | • **Yes:** if people selected from the sampled facilities (e.g. hospital outpatient department) are representative of the people in general who go to those facilities<br>• **No:** if people selected from the sampled facilities are not representative of the people in general who go to those facilities<br>• **Unclear:** if unclear whether people selected from the sampled facilities are representative of the people in general who go to those facilities |
| Were study participants sampled in a manner that was adequately described? | • **Yes:** if sampling scheme can be understood in sufficient detail from the study description<br>• **No:** if the sampling scheme is not adequately described<br>• **Unclear:** if the sampling scheme description is unclear |
| Were the study subjects and the setting described in detail? | • **Yes:** if study subjects and the setting described in detail (e.g. data could be extracted for these points)<br>• **No:** if the study subjects and the setting were not described in sufficient detail<br>• **Unclear:** if the description of the study subjects and the setting is unclear |
| Was the condition measured in a standard, reliable way for all participants? | • **Yes:** if the Ag-RDT was performed and interpreted in a standard, reliable way for all participants (e.g. explicitly stated as per manufacturer's instructions, or using the same procedures for each participant)<br>• **No:** if the Ag-RDT was performed and interpreted in different ways for different participants<br>• **Unclear:** if the description of how the Ag-RDT was conducted and interpretated is unclear |
| Was there appropriate statistical analysis? | • **Yes:** if the statistical analyses are appropriate for the reported outcomes<br>• **No:** if the statistical analyses are not appropriate for the reported outcomes, or appropriate statistical analyses not conducted<br>• **Not applicable:** if study did not include data requiring statistical analyses e.g. studies reporting outcomes like sensitivity, test positivity, or purely operational outcomes |

**Grading:** Yes = low risk; No = high risk; Unclear = unclear risk.

### Supplementary Table 4: Detailed individual study results

| First author, reference | Country | Study details | Brand of test and name of test; swab type; test performer | Sample size (n) | Sensitivity % (95% CI) | Specificity % (95% CI) | Other outcomes and operational findings | Author conclusions |
|---|---|---|---|---|---|---|---|---|
| **Mass screening** | | | | | | | | |
| Iglὀi[1] | The Netherlands | Largest drive-through testing site in Rotterdam, by appointment only. Eligibility for free test if symptomatic or close contact of confirmed case. 18 years and older. | SD Biosensor SARS-CoV-2 Rapid Antigen Test; NP swab; Unknown | 970 | 84.9 (79.1–89.4) | 99.5 (98.7–99.8) | 95% of strong positive samples appeared in <5 minutes. | "Screening based on Ag RDT alone in this population would have a high sensitivity for ruling out infectious individuals." |

| | | | | | | | | |
|---|---|---|---|---|---|---|---|---|
| Pilarowski[2] | USA | Walk-up free testing offered over 3 days to asymptomatic or symptomatic adults at a plaza in a neighbourhood of San Francisco known to have a high prevalence of SARS-CoV-2 infection. | Abbott BinaxNOW COVID-19 Ag Card; nasal swab; lab staff | 878 | 57.7<br><br>(36.9–76.6) | 100<br><br>(99.6–100) | The manufacturer's suggestion to score any visible bands as positive led to excessive false-positives (64%). Optimal performance occurred when the bands were scored as positive if they extended across the full width of the strip, irrespective of the intensity of the band. | "Binax-CoV2 test should not be limited to symptomatic testing alone. Limiting use of Binax-CoV2 to symptomatic individuals would have missed nearly half of the SARS-CoV-2 infections in the current study." |
| Pollock[3] | USA | Drive-through community testing site outside a hospital in Lawrence, Massachusetts, accommodating symptomatic or asymptomatic children and adults from the surrounding area. | Abbott BinaxNOW COVID-19 Ag Card; nasal swab; lab staff | 2482 | Asx adults: 70.2 (56.6–81.6)<br><br>Asx children: 65.4 (55.6–74.4) | Asx adults: 99.6 (98.9–99.9)<br><br>Asx children: 99.0 (98.0–99.6) | In 30 specimens run at temperatures below manufacturer's recommendations, sensitivity was 66.7% and specificity was 95.2%. Excellent inter-operator agreement. Skilled lab staff can perform | Sensitivity decreased with increasing Ct values. |

| | | | | | | | | |
|---|---|---|---|---|---|---|---|---|
| | | | | | | | and read 20 tests per hour. | |
| Sood[4] | USA | Families with children <18 years at a walk-up testing site in Los Angeles County for those seeking COVID-19 testing because they were symptomatic or had been in contact with a case. Results from children were read by two trained staff. | Abbott BinaxNOW COVID-19 Ag Card; Nasal swab; Other trained personnel | 779 | 56.2 (49.5–62.8) | NA | Positive concordance was higher among children with lower Ct values on the RT-PCR test, including in asymptomatic children. | "Rapid antigen testing can identify most COVID infections in children with viral load levels likely to be infectious and serial rapid testing may help compensate for limited sensitivity in early infection." |
| Pilarowski[5] | USA | Community workers conducted door-to-door mobilization near the testing site four days before testing. Testing was conducted at a plaza under tents in San Francisco, in a setting of ongoing community transmission, | Abbott BinaxNOW COVID-19 Ag Card; Nasal swab; Lab staff | 3302 | 89.0 (84.3–92.7) | 99.9 (99.7–100) | Among symptomatic persons with positive Ag-RDT, median time from symptom onset to isolation was 3 days (IQR: 2–5 days). For those with positive Ag-RDT, median time from onsite registration to | "Integration of rapid antigen testing within community-based test and respond initiatives could contribute to reduced transmission via several mechanisms… our low-barrier testing |

| | | | | | | | | |
|---|---|---|---|---|---|---|---|---|
| | | predominantly among Latinx persons. | | | | | electronic results notification was 62 min. With 3 tents and 13 total staff, 100 persons tested/hour. | model… linked with supportive follow-up services could identify more infectious persons faster, decrease the time to isolation, and interrupt transmission chains.” |
| Shah[6] | USA | Community SARS-CoV-2 testing site in Oshkosh, Wisconsin, during an ongoing mass surge testing campaign. | Abbott BinaxNOW COVID-19 Ag Card; nasal swab; self-taken sample observed by trained staff | 2110 | 77.2<br>(72.4–81.6) | 99.6<br>(99.2–99.8) | The BinaxNOW test exhibits minimal user error and high concordance (98.9%) when repeated at the same encounter offering low yield for capturing additional COVID-19 cases. | “In certain settings with high prevalence and limited resources, it may also be reasonable to forego RT-PCR confirmation in asymptomatic BinaxNOW positive individuals." |
| Pollock[7] | USA | Drive-through community testing site outside a hospital in Lawrence, Massachusetts, testing asymptomatic adults | Access Bio CareStart COVID-19 antigen; nasal swab; lab staff | 1498 | 50<br>(41.0–59.0) | 99.1<br>(98.3–99.6) | Median interval between sample collection and test initiation was 31 minutes (range 12-103 minutes). Excellent inter-operator agreement. Additional training of personnel warranted | Higher sensitivity in people with Ct<25. Observed specificity lower than stated in instructions for use. |

| | | | | | | | | |
|---|---|---|---|---|---|---|---|---|
| | | | | | | | to mitigate test failures | |
| Seitz[8] | Austria | Testing of healthy citizens of all ages invited for voluntary COVID-19 screening. Those with positive NP swab Ag-RDT and had gargle sample RT-PCR were invited to participate. | Xiamen Zhongsheng Langjie Biotechnology; Saliva; Healthcare worker/clinician | 40 | 44.4 | 100.0 | Self-testing with Ag-RDT on saliva allows cheap, simple, and fast testing for SARS-CoV-2. | "Our data suggest that even individuals with high virus load may not be detected by saliva RAT. A negative saliva RAT test cannot confirm the absence of SARS-COV, a confirmation with RT-PCR is needed." |
| Alemany[9] | Spain | Retrospective analysis of frozen samples from individuals attending mass screening campaigns, population was asymptomatic, symptomatic, and contacts exposed to symptomatic | Abbott Panbio COVID-19 Ag Rapid Test; Nasal Swab, Unknown | 487 | 79.5 | NA | Internal validation showed no relevant differences between tests performed on fresh samples using the Abbot test Kit buffer and 1:3 dilutions of the Kit buffer and frozen specimens stored on transport media | In high prevalence settings, Ag-RDTs are useful for screening asymptomatic individuals for COVID-19; high sensitivity, therefore suitable for creating safe environments in time-limited social |

| | | | | | | | | |
|---|---|---|---|---|---|---|---|---|
| | | | | | | | | activities with high-risk of transmission. |
| Green[10] | UK | Analysis of spatial and social inequalities of rapid antigen testing in city-wide testing conducted in Liverpool. Participants were asymptomatic city residents, with recruitment emphasis on hot-spot neighbourhoods, students, and other high-risk groups. | Unknown; Unknown; Unknown | 399,603 | NA | NA | Testing rates were higher in females and working age-adults. Uptake and repeat testing were low in BAME groups and in the most deprived areas, locations further from test sites, and in populations with less confidence in using internet technology. These groups were more likely to have positive test results. | “Provision of free and voluntary asymptomatic community testing is affected by substantial social and spatial inequalities, typical of the ‘inverse care’ law but with a distinctive digital exclusion factor consistent with the digitally intensive means of accessing testing.” |

| | | | | | | | | |
|---|---|---|---|---|---|---|---|---|
| Frnda[11] | Slovakia | Nationwide mass testing of population ages 10–65. In the trial period, four districts with high COVID-19 incidence were included; one week later, the entire country was tested. | Abbott, Biosensor Standard, RapiGEN, Unknown; Unknown | 140,945 (trial in four districts); 3,625,332 (nationwide) | NA | NA | Positivity rates were 3.97% in 4 trial districts and 1.06% nationwide. Nationwide testing occurred over 2 days. In very low prevalence regions, test credibility is questionable due to (hard-to-estimate) false-positive results. | "In regions with a very low expected prevalence, mass testing is ineffective and unworthy of its financial and human capital costs… However, in general, nationwide testing by antigen tests has identified many potentially infected individuals within the population that would not be discovered, yet they would be involved in the spread of the virus, causing COVID-19." |
| Pavelka[12] | Slovakia | Phased evaluation of nationwide testing among entire Slovakian population older than 10 years; >80% of age-eligible population tested in each round | SD Biosensor STANDARD Q COVID-19 Ag Test; NP swab; took sample: HCW/clinician, ran test: other trained personnel | 5,276,832 | NA | NA | Test positivity - pilot: 3.91%, round 1: 1.01%, round 2: 0.62%. Getting enough trained medical personnel to take NP samples was noted as a challenge; difficult to untangle effect of mass testing from stricter public health measures. | "The combination of nationwide restrictions and mass testing with quarantining of household contacts of test positives rapidly reduced the prevalence of infectious residents in Slovakia." |

| | | | | | | | | |
|---|---|---|---|---|---|---|---|---|
| Colavita[13] | Italy | Voluntary on-site testing of travellers returning from high-incidence foreign countries, as well as from the Sardinia region at points of entry (airports and ports) in the Lazio region. | SD Biosensor STANDARD F COVID-19 Ag Fluorescence Immunoassay; NP swab; Healthcare worker (for both) | 73,643 | NA | NA | Test positivity was 1.6%; In low prevalence setting, 40.5% samples confirmed with RT-PCR of which 60% were false positive. | “The data show that the probability of (RT-PCR) confirmation of positive results by STANDARD F COVID-19 Ag FIA is directly related to the cut-off index values from this semi-qualitative rapid test.” |
| **Targeted screening** | | | | | | | | |

| | | | | | | | | |
|---|---|---|---|---|---|---|---|---|
| Okoye[14] | USA | Mass screening of asymptomatic young adult university students in Salt Lake City | Abbott BinaxNOW COVID-19 Ag Card; nasal swab; self-taken sample observed by trained non-medical personnel | 2645 | 53.3<br><br>(39.1–67.1) | 100<br><br>(99.9–100) | Kappa value of all positivity was 0.69 (95% CI 0.57–0.82). Test was able to be performed successfully at the point of care by trained nonmedical personnel, with a relatively low rate of invalid results (0.1%) | "Despite its relatively low analytical sensitivity, BinaxNOW may still be beneficial for surveillance testing in selected settings where testing resources are limited, especially when weighed against the alternative of no screening testing" |

| | | | | | | | | |
|---|---|---|---|---|---|---|---|---|
| Boum[15] | Cameroon | Adult symptomatic suspected cases or already on treatment; asymptomatic adults presenting for voluntary screening or referred through contact tracing in 8 sites in Yaoundé, Cameroon | SD Biosensor Ag-RDT; NP swabs; other trained personnel | 1195 | 59.0<br><br>(53.0–65.0) | 94.0<br><br>(88.0–97.0) | NA | Ag-RDTs have been a critical foundation of Cameroon's national testing strategy, allowing decentralized and mobile testing in public places. Ag-RDTs are well-suited to low-resource environments, especially settings where PCR testing has restricted availability. |

| | | | | | | | | |
|---|---|---|---|---|---|---|---|---|
| James[16] | USA | Mandatory screening of all healthcare workers providing patient care in an acute care hospital in Arkansas. 95% asymptomatic | Abbott BinaxNOW COVID-19 Ag Card; nasal swab; clinician or lab staff | 2339 | 56.6<br><br>(48.7–64.5) | 99.9<br><br>(99.7–100) | Sensitivity 83.3% in symptomatic and 51.6% in asymptomatic people. | “Despite the lower sensitivity... these tests could be strategically paired with rRT-PCR testing to immediately identify and isolate persons potentially at higher risk of transmitting the infection while rRT-PCR results are pending.” |

| | | | | | | | | |
|---|---|---|---|---|---|---|---|---|
| Yokota[17] | Japan | All arrivals at three international airports were tested for COVID-19. Each traveller first underwent an Ag-RDT. Those with “indeterminate” test results (i.e., within positive and negative thresholds of 4.0 pg/mL and 0.67 pg/mL) had to undergo additional molecular testing. | Fujirebio; Lumipulse SARS-CoV-2 Ag**; saliva; lab staff | 88,924 | NA | NA | Positivity rate was 0.29%. 513 results were within the indeterminate threshold, of which 34 were confirmed positive. The two-step strategy led to a 95% reduction in molecular test use compared to universal deployment, saving time and resources. | “Two-step testing by CLEIA followed by NAAT is effective in real-world situations, especially when combined with appropriately timed pre-departure testing and/or with quarantine optimized with repeat testing.” |

| | | | | | | | | |
|---|---|---|---|---|---|---|---|---|
| Gili [18] | Italy | Unselected cohort of swabs collected in schools, prisons, elderly care homes, and from hospital healthcare worker surveillance programs in Umbria was used. | Fujirebio; Lumipulse SARS-CoV-2 Ag**; NP swab; Lab staff | 1738 | 100 | 92.1 | Tests were run on a multiplex reader, so process was completely automated. Up to 120 samples/hour may be run. | "Lumipulse® SARS-CoV-2 antigen assay, compared with RT-PCR from nasopharyngeal swabs, showed an excellent NPV for the presence of SARS-CoV-2 infection both in high- and low-prevalence scenarios, supporting the use of this assay in selected high-risk communities and for community and population screening purposes." |

| | | | | | | | | |
|---|---|---|---|---|---|---|---|---|
| Pray[19] | USA | Adult symptomatic (21%) or asymptomatic (79%) university students and staff at two universities in Wisconsin | Quidel Sofia SARS Antigen Fluorescent Immunoassay (FIA); nasal swab; trained healthcare workers or self-test observed by trained healthcare workers | 871 | 41.2 (18.4–67.1) | 98.4 (97.3–99.1) | All false-negative results from symptomatic participants were from specimens collected <5 days after onset of symptoms (median = 2 days). | Authors recommend confirmatory testing with an FDA-authorized NAAT, such as RT-PCR, following negative antigen test results in symptomatic persons to minimize the impact of false negatives. |

| | | | | | | | | |
|---|---|---|---|---|---|---|---|---|
| Betancourt[20] | USA | In a university dorm of Arizona, pre-screening of wastewater was done with RT-PCR; If positive, testing of entire dorm with Ag-RDT was done; Additional symptomatic students also tested | Quidel Sofia SARS Antigen Fluorescent Immunoassay; Nasal Swab; Unknown | 311 | NA | NA | 50% effectiveness wherein Ag-RDT identified 1 person who was also RT-PCR+, missed 1 person who was RT-PCR indeterminate (later positive); Waste water surveillance + targeted testing seems effective and acceptable. | "The combined strategy of utilizing WBE with targeted clinical testing was critical in COVID-19 containment. WBE strategies averted potential transmission from at least three students, which allowed the university to remain open, and even establish limited in-person classes for students." |

| | | | | | | | | |
|---|---|---|---|---|---|---|---|---|
| Moreno[21] | USA | Asymptomatic students and staff affiliated with two university athletics programmes were tested daily with Ag-RDT, with positive results confirmed using RT-PCR | Quidel Sofia SARS Antigen Fluorescent Immunoassay; NP, nasal swab; took sample: self sample (nurse supervised) | Unknown | NA | NA | Antigen testing on the competition dates failed to identify the index case, who may have been infectious and exposed other athletes. | "…serial antigen testing as a control strategy may have limited sensitivity for detecting early asymptomatic infections" |

| | | | | | | | | |
|---|---|---|---|---|---|---|---|---|
| Herrera*[22] | USA | Testing of HCWs at ambulatory centres, who had come in contact with confirmed cases, not sooner than 4-5 days post exposure if asymptomatic, or sooner if symptomatic | Unknown: test brand/name, sample and who took sample and ran test | 497 | NA | NA | Test positivity: symptomatics = 11%, asymptomatics = 2%. No asymptomatic antigen-negative HCWs developed symptoms or had evidence of transmitting SARS-CoV-2. Due to low RT-PCR positivity rate, samples were pooled to further conserve PCR capacity. | "…a rapid antigen testing process is an effective strategy that relieves anxiety, decrease resource utilization, and facilitates return to work" |

| | | | | | | | | |
|---|---|---|---|---|---|---|---|---|
| Cerutti[23] | Italy | Travellers (mean age 35.9 years) returning home from European high-risk countries (Croatia, Spain and Malta) in August 2020 | SD Biosensor STANDARD Q COVID-19 Ag Test; NP swab; took sample and ran test: unknown | 145 | 40 | 100 | NA | "Sensitivity, specificity, accuracy, negative and predictive values were consistent with the use of the test to mass-screening for SARS-CoV-2 surveillance." |

| | | | | | | | | |
|---|---|---|---|---|---|---|---|---|
| Peto[24] | UK | Nationwide phased evaluations of Ag-RDTs: retrospective analysis of patient samples from a secondary healthcare setting; drive-through community testing; community field evaluations (secondary healthcare [hospital] setting, armed forces, Public Health England staff, school children; regional COVID-19 testing centres) | Innova SARS-CoV-2 Antigen Rapid Qualitative Test; Nasal, NP swab, oropharyngeal swab; self collected sample, test ran by lab staff | 6954 | NA | 99.68 | Poor transfer of the liquid within the device from the reservoir onto the test strip was the most common reason for kit failure; In field testing, performance was dependent on the test operator. Individuals who had read a protocol immediately prior to self-sampling did not perform as well as individuals with hands-on training. | Ag-RDTs are promising for mass population testing. The Innova lateral flow device shows good viral antigen detection/sensitivity with excellent specificity, although kit failure rates and the impact of training are potential issues. |
| **Healthcare entry testing** | | | | | | | | |

| | | | | | | | | |
|---|---|---|---|---|---|---|---|---|
| Regev-Yochay*[25] | Israel | Screening of asymptomatic patients upon hospitalization and a cohort of HCWs following SARS-CoV-2 exposure at the largest tertiary hospital in Israel | Nowcheck COVID-19 Ag test (Bionote), Panbio COVID-19 Ag rapid test, (Abbott), BD Veritor (BD), GenBody COVID-19 Ag (GenBody), STANDARD Q COVID-19 (SD-Biosensor); NP swab; took sample: other trained personnel, ran test: lab staff | 1548 | 65.9 | 99.8 | NA | “Ag-RDT can be used as a decision support tool in various clinical settings and play a major role in early detection of SARS-CoV-2” |

| | | | | | | | | |
|---|---|---|---|---|---|---|---|---|
| Rottensreich [26] | Israel | Testing of asymptomatic pregnant women admitted for delivery in a university-affiliated hospital | Bionote NowCheck COVID-19 Ag Test; NP swab; unknown | 1326 | 55.6<br><br>(21.2–86.3) | 100<br><br>(99.7–100) | NA | “A universal testing approach using RDT among women admitted for delivery may allow timely determination of COVID-19 status that will guide the utilization of proper protection measures and inform neonatal care.” |

| | | | | | | | | |
|---|---|---|---|---|---|---|---|---|
| Tripathy[27] | India | Preoperative screening of all asymptomatic patients undergoing elective ophthalmic surgeries; testing of staff with presumptive COVID-19; back to work policy of in-house staff in a tertiary eye hospital in Odisha | SD Biosensor STANDARD Q COVID-19 Ag Test; NP swab; lab staff | 311 | NA | NA | Mandatory Ag-RDT testing appeared to increase surgery patients' feeling of safety around seeking medical care during the pandemic. | Ag-RDTs “may be considered routinely for indication-based preoperative screening of asymptomatic patients, and for on-campus screening, contact tracing and implementation of [back-to-work] policies” for healthcare workers. |

| | | | | | | | | |
|---|---|---|---|---|---|---|---|---|
| Turcato[28] | Italy | Initial screening of all patients presenting at the hospital emergency department irrespective of health condition | SD Biosensor Standard Q COVID-19 Ag Test; unknown; unknown | 3410 | 80.3 (74.9–85.4) | 99.1 (98.6–99.3) | Use of Ag-RDTs for screening of symptomatic and asymptomatic patients upon arrival in the ED can improve the overall management of the infectious risk, with a net clinical benefit. | Based on experience in their hospital emergency department, the authors recommend the use of Ag-RDTs for infectious risk assessment during triage in emergency departments. |

| | | | | | | | | |
|---|---|---|---|---|---|---|---|---|
| Van Honacker[29] | Belgium | Screening for all hospital emergency department patients with an indication for hospital admission of whom there was no recent positive SARS-CoV-2 rRT-PCR result available (<8 weeks) | SD Biosensor SARS-CoV-2 Rapid Antigen Test; NP swab; unknown | 4195 | 54.2 | 99.7 | Incorrect reading of test cassette caused false positive in 3 out of 12 results. | Despite low sensitivity, acceptable performance of Ag-RDTs for ED patients as most infectious individuals were detected; Ag-RDTs can be used as a rapid screening tool in high prevalence settings but cannot replace RT-PCR in ED. |

| | | | | | | | | |
|---|---|---|---|---|---|---|---|---|
| Dalal[30] | India | Exposed or symptomatic healthcare workers and all patients visiting hospital outpatient department underwent Ag-RDT as initial screening before gastrointestinal endoscopy. | SD Biosensor Standard Q COVID-19 Ag Test; unknown; unknown | Unknown | NA | NA | 3.8% healthcare workers performing endoscopic procedures were diagnosed with COVID-19; only 1 procedure was done on an Ag-RDT+ person; Ag-RDT can be used for emergency situations in the endoscopy unit. | "Our study shows that the RAT for COVID-19 is an easy, economical, and convenient screening tool for patients undergoing GI endoscopic procedures" |
| **At-home testing** | | | | | | | | |

| | | | | | | | | |
|---|---|---|---|---|---|---|---|---|
| Martin[31] | UK | Online acceptability survey of a pilot study of at-home testing to replace self-isolation. Participants were asymptomatic contacts of COVID-19 cases who consented to rapid antigen self-testing for 7 days. Comparisons were made to eligible adults who isolated, and eligible adults who did not consent to participate. | Unknown; Unknown; Self | 319 | NA | NA | 62% accepted daily testing. Acceptability was lower in minority groups. 88% were confident they performed the test correctly. Common issues included (i) Internet/technology access, (ii) tests being unpleasant, and (iii) unclear instructions. | "Daily testing is potentially acceptable, may facilitate sharing contact details of close contacts among those who test positive for COVID-19, and promote adherence to self-isolation." "The impact of receiving a negative test on behaviour remains a risk that needs to be monitored and mitigated by appropriate messaging." |

| | | | | | | | | |
|---|---|---|---|---|---|---|---|---|
| Downs[32] | England | Study of home-based testing of asymptomatic healthcare workers and support staff. | Innova; Unknown; Nasal swab; Self | 8657 | NA | NA | Positivity rate was 0.7% (95% CI: 0.01–0.06); 1.0% of tests were invalid. Staff found testing kits easy to use. Kit failure rates and false positive results in this population were low enough to support their widespread use in asymptomatic population. | “Use of LFDs identified asymptomatic SARS-CoV-2 infections that would not otherwise have been detected... Although individuals with low viral loads may be missed, with regular serial testing at least twice per week, those with early infection and rising viral loads are likely to be identified.” |

| | | | | | | | | |
|---|---|---|---|---|---|---|---|---|
| Hoehl*[33] | Germany | Teachers from primary and secondary schools in three school districts tested themselves at home repeatedly for 7 weeks to reduce transmission in school. | R-Biopharm RIDA QUICK SARS-CoV-2 Antigen test; Nasal Swab; Self | 711 | NA | NA | Test Positivity was 0.19%; Ag-RDT can be performed satisfactorily unsupervised at-home by the participant; medical/technical assistance through hotline number should be provided; participants found it reassuring. | "High-frequency, self-performed and repeat rapid antigen tests in a school setting can detect COVID-19 in a timely manner and prevent onward transmission; most beneficial in a high incidence setting and mild/atypical symptoms are present." |

| | | | | | | | | |
|---|---|---|---|---|---|---|---|---|
| Love*[34] | UK | Testing of asymptomatic adult contacts exposed to COVID-19 case within the preceding 48 hours using serial, self-administered RATs as an alternative to self-isolation for 7 days post exposure. | Innova; nasal swab; took sample and ran test: self | 882 | NA | NA | Test positivity: 17.9%. Most common motivation for consenting to daily self-testing were a duty to take part (33.8%), the assurance of daily testing (30.4%) and not wanting to self-isolate (26.0%). | "This study shows a high acceptability, compliance and positivity rates when using self-administered LFDs among contacts of confirmed COVID-19 cases" |
| **Surveillance** | | | | | | | | |

| | | | | | | | | |
|---|---|---|---|---|---|---|---|---|
| Smith[35] | USA | All on-campus students and employees of the University of Illinois at Urbana-Champaign are required to submit saliva for RT-qPCR testing every 2-4 days as part of the SHIELD campus surveillance testing programme. Participants were selected from this programme to provide daily samples for RT-qPCR, Ag-RDT and live virus culture. | Quidel Sofia SARS Antigen Fluorescent Immunoassay; Nasal swab; swabs self-collected under supervision, test run by lab staff | 43 | ~90% for daily screening while individual is viral culture positive | NA | Sensitivity of Ag-RDT varies over the course of SARS-CoV-2 infection. The Ag-RDT peaks in sensitivity during the period in which live virus can be detected in nasal swabs. | “All tests showed >98% sensitivity for identifying infected individuals if used at least every 3 days. Daily screening using antigen tests can achieve approximately 90% sensitivity for identifying infected individuals while they are viral culture positive.” |

| | | | | | | | | |
|---|---|---|---|---|---|---|---|---|
| Kotsiou[36] | Greece | Two passive surveillance programmes of entire population in Volos using Ag-RDT before and during lockdown; mixture of symptomatic and asymptomatic. | VivaChek Biotech VivaDiag SARS-CoV-2 Antigen Rapid Test; nasal swab; took sample: HCW/clinician, ran test: lab staff | 1054 (pre-lockdown screening); 462 (lockdown group) | NA | NA | Test positivity: 8% (pre-lockdown group); 4.7% (during lockdown group). Positive participants were more likely to work in the catering/food sector than negative participants before the lockdown. Lockdown restrictions halved the new cases. | This study “highlight(s) the crucial role of community-based screening with rapid antigen testing to evaluate the potential modes of community transmission and the impact of infection control strategies” |

| | | | | | | | | |
|---|---|---|---|---|---|---|---|---|
| Winkel[37] | The Netherlands | Longitudinal cohort study testing asymptomatic football players and staff members of professional football clubs; age range 16 to 80 years. | Abbott PanBio COVID-19 Ag Rapid Test; NP swab; took sample and ran test: other trained personnel | 824 | 85.71 (67.3 to 96.0) | 100 (99.8–100) | False-negative results were mostly observed in late phase of infection (on average 2 weeks after first positive test result). | “the Panbio COVID-19 Ag rapid test is able to identify early SARS-CoV-2 infections in asymptomatic individuals and can be used in targeted screening strategies for detection of SARS-CoV-2 infection in asymptomatic individuals. Caution is advised in situations where high test sensitivity is critical” |

| | | | | | | | | |
|---|---|---|---|---|---|---|---|---|
| Kriemler [38] | Switzerland | Asymptomatic and symptomatic students and teachers from 14 primary and secondary schools in Zurich underwent testing with Ag-RDT and PCR on two days one week apart | SD Biosensor, STANDARD Q COVID-19 Ag Test; buccal swab; took sample and ran test: unknown | 707 | NA | 99.4 | NA | "Specificity of the RDT was within the lower boundary of performance and needs further evaluation for its use in schools" |
| **Prevalence survey** | | | | | | | | |

| | | | | | | | | |
|---|---|---|---|---|---|---|---|---|
| Babu [39] | India | Cross-sectional survey of symptomatic and asymptomatic adults across Karnataka, participants were tested using ELISA, Ag-RDT and RT-PCR | SD Biosensor STANDARD Q COVID-19 Ag Test; NP swab, oropharyngeal swab; took sample and ran test: unknown | 8673 | Symptomatic = 68.0, Asymptomatic = 46.9 | NA | Significant predictive variables for active infection: headache, chest pain, wheezing, rhinorrhoea, cough, sore throat, muscle ache, fatigue, chills, and fever; additional predictive variables: hospital outpatient attendance and contact with COVID-19 patients. | The combined estimates (of past SARS-CoV-2 infection from antibody tests and active infection from Ag-RDT/RT-PCR) could lead to timely, informed and evidence-based public health responses. |

Key: *Preprint; **Multiplex test - all other tests standalone strips.

Ag-RDT/RAT, antigen-detecting rapid diagnostic test; asx, asymptomatic; CLEIA, chemiluminescence enzyme immunoassay; Ct, cycle threshold; ED, emergency department; FDA, Food and Drug Administration; GI, gastrointestinal; HCW, healthcare worker; LFD, lateral flow device; NAAT, nucleic acid amplification test; NP, nasopharyngeal; NPV, negative predictive value; PCR, polymerase chain reaction; RT-PCR, reverse transcription polymerase chain reaction; rRT-PCR, real-time reverse transcription polymerase chain reaction; RT-qPCR, quantitative reverse transcription polymerase chain reaction; WBE, wastewater-based epidemiology.
